# Patient perspectives on cardioprotective medication during breast cancer treatment

**DOI:** 10.64898/2026.06.29.26356895

**Authors:** Lauren Houston, Nusayba A. Bagegni, Mei Ling Yap, Elgene Lim, Bruce Neal, Anita Deswal, Joshua D. Mitchell, Clare Arnott, Sang Gune K. Yoo

**Author notes:** **Corresponding Author:** Sang Gune K. Yoo, MD, Assistant Professor of Cardiology, Clinical Cancer Prevention, UT MD Anderson, 1515 Holcombe Blvd, Unit 1451, Houston, TX, 77004.

## Abstract

**Background:** Cardiotoxicity remains a key concern of HER2-directed therapies, with no proven prevention strategies. The success of cardioprotective interventions will depend not only on efficacy, but on patient acceptability, an underexplored determinant of trial participation and clinical implementation.

**Objectives:** To evaluate willingness to take cardioprotective medication and identify factors influencing decision-making among individuals with HER2-positive breast cancer.

**Methods:** We conducted a cross-sectional online survey of adults with HER2-positive breast cancer in Australia and the United States. The survey assessed willingness, beliefs regarding benefits and risks, and treatment preferences. Multivariable logistic regression examined associations between clinical and demographic characteristics and willingness.

**Results:** Among 74 respondents (Australia n=24; United States n=50), 74.3% reported being likely or very likely to take cardioprotective medication. Physician recommendation emerged as a dominant driver (79.1%). While most participants valued long-term cardiovascular health (72.9%), uncertainty regarding benefit was common (60.4%). Cancer-related outcomes were prioritized over cardiovascular outcomes. Participants demonstrated flexibility regarding treatment burden, including willingness to take multiple medications and continue therapy long-term. No demographic or clinical predictors of willingness were identified. Perceived acceptability, appropriateness, and feasibility were consistently high.

**Conclusions:** Willingness to adopt cardioprotective strategies is high but conditional, shaped by cancer priorities, clinician endorsement, and uncertainty regarding benefit. These findings highlight patient acceptability as a critical, and often overlooked, determinant of successful trial participation and downstream clinical implementation in cardio-oncology.

## Introduction

Breast cancer is the most common cancer worldwide and the leading cause of cancer-related deaths among women.^1^ Approximately 10–15% of breast cancers overexpress the human epidermal growth factor receptor 2 (HER2), a subtype historically associated with accelerated cell proliferation and worse clinical outcomes.^2,3^ The development of HER2-directed therapies, including monoclonal antibodies, has transformed outcomes for patients with this breast cancer subtype. However, these therapies are associated with an increased risk of cardiotoxicity, including both asymptomatic left ventricular dysfunction and clinical heart failure.^4–8^ When a clinically significant decline in left ventricular ejection fraction is detected, interruption or cessation of HER2-directed therapy may be required, potentially compromising the anti-cancer and survival benefits these therapies provide. Treatment interruption due to cardiotoxicity affects approximately 15-20% of patients and has been associated with worse oncologic outcomes in observational studies.^9,10^

In the context of improving cancer survival, cardiovascular disease has emerged as a leading cause of morbidity and mortality among breast cancer survivors.^11^ Despite this, no proven pharmacologic strategy exists to prevent or reduce the risk of cardiotoxicity in patients receiving HER2-directed therapy. Although there is ongoing interest in reducing cardiotoxicity in this setting, the evidence base remains inadequate and clinical trials have been inconclusive.^12^ Patient acceptability of cardioprotective strategies remains poorly characterized, yet is a critical determinant of both trial participation and subsequent clinical implementation. Understanding patient perspectives is therefore essential to inform the design and feasibility of future cardiotoxicity prevention trials and their translation into clinical practice.

This study examines patient perspectives on the acceptability of cardioprotective medication through a survey of individuals with a history of HER2-positive breast cancer in Australia and the United States (US). Specifically, we sought to characterize patient’s willingness to accept additional cardioprotective medication and explore potential factors that influence decision-making.

## Methods

### Setting

We conducted a cross-sectional, semi-quantitative online survey among individuals with HER2-positive breast cancer in Australia and the US. The survey was administered using REDCap (Research Electronic Data Capture), a secure, web-based platform used to collect and manage study data. Ethics approval was obtained from St Vincent’s Hospital Sydney Human Research Ethics Committee (2022/ETH00514) in Australia and Washington University School of Medicine/Siteman Cancer Center Institutional Review Board (HRPO# 202508209) in the US.

### Participants

Eligible participants were adults (≥18 years) with HER2-positive breast cancer who were commencing, undergoing, or had previously completed HER2-directed therapy at the time of recruitment (no restriction on time since completion) and were able to read and understand English. Potentially eligible individuals were identified through screening clinic lists and routine outpatient appointments across two breast oncology clinics in Australia and two breast oncology and cardio-oncology clinics in the US. This represents a convenience sample of patients attending participating clinics. All demographic and clinical characteristics were self-reported. This included treatment history (early-stage vs metastatic), treatment duration (categorical) and treatment interruption (yes/no/don’t know), comorbidities, and medication use, and responses were not independently verified.

### Survey design and development

The reporting of survey design and results followed the Checklist for Reporting Results of E-Internet Surveys (CHERRIES).^13^ A self-administered, semi-quantitative survey was developed by a multidisciplinary research team with expertise in cardiology and oncology. The survey was informed by a systematic review and meta-analysis of heart failure therapies for the prevention of cardiotoxicity associated with HER2-directed therapy.^12^ The questionnaire included three domains: (1) participant demographics, (2) clinical characteristics, and (3) willingness to take additional cardioprotective medication. The core survey contained a total of 11 items; one additional item was included in the Australian survey (total n = 12), and two additional items in the US survey (total n = 13) relating to race/ethnicity. Four items incorporated adaptive logic, with branching determining the total number of questions presented based on individual survey responses. An additional two items were included in the US survey to assess beliefs about benefits, risks, and treatment burden, as well as acceptability of cardioprotective medication during cancer treatment. These items were informed by established implementation outcome frameworks, including the Acceptability of Intervention Measure, Intervention Appropriateness Measure, and feasibility of Intervention Measure.^14^ Survey items included multiple-choice questions (single and multiple selection), binary items (yes/no), open-ended free-text responses, and five-point Likert-scale items, including grouped Likert statements. Responses to all items were voluntary, and respondents could skip items or select non-responses, such as “not applicable” or “don’t know”. Survey respondents were provided with standardized explanatory text within the survey describing cardiotoxicity risk and the potential role of cardioprotective medications. The survey was pilot tested in a convenience sample of five individuals to assess clarity, readability, and question flow, with minor refinements made prior to dissemination. The Australian and US survey instruments are provided in the **Supplementary Appendix 1 and 2.**

### Survey administration

The survey was opened in Australia from June 2022 to July 2024 and in the US from September 2024 to April 2026. Participants were recruited using a combination of in-clinic and remote approaches. In clinic settings, potentially eligible individuals were approached by trained research staff and provided with a QR code or the survey URL to complete on their own device. In the US cohort, a paper version or tablet was available on request. For remote recruitment, participants were contacted via telephone or email and provided with the survey URL. The total number of individuals approached and invited to participate was not systematically recorded due to the combined use of in-clinic and remote recruitment. Everyone was invited to complete the survey once to minimize duplicate responses. Completion of the survey following an electronic consent statement constituted implied consent. The survey took up to 15 minutes to complete.

### Data analysis

All available responses, including partially completed surveys, were included in the analysis to minimize potential bias associated with differential item completion. Data were analyzed using R (version 4.5.2; R Foundation for Statistical Computing, Vienna, Austria) in RStudio (Posit Software, Boston, MA, USA).

Descriptive statistics summarized participant characteristics and survey responses, presented as counts and percentages. Percentages were calculated using available data for each variable, and denominators may very due to missing responses or survey skip logic. Percentages represent column percentages unless otherwise specified. Missing data were not imputed. Categorical variables were compared between countries (Australia vs US) as the primary analysis using Fisher’s exact test, given small cell counts. Given clinically meaningful differences between early-stage and metastatic disease, cancer stage-stratified analyses were conducted as a secondary exploratory analysis using the same approach. A two-sided p<0.05 was considered to indicate statistical significance. For ‘select all that apply’ items, responses were summarised for each option. Likert-scale responses were analyzed across all categories and, where appropriate, collapsed into three groups: disagree (completely disagree/disagree), neutral (neither agree nor disagree), and agree (agree/completely agree) for summary figures, presented as stacked bar charts. Free text responses were analyzed using content analysis.^15^

Multivariable logistic regression was used to explore associations between demographic and clinical characteristics and willingness to take cardioprotective medication. Willingness was treated as a binary outcome (likely/very likely vs neutral/unlikely/very unlikely). Two separate multivariable models including pre-specified demographic and clinical variables were used to estimate adjusted odds ratios (ORs) with 95% confidence intervals (CIs), with the number of variables limited to reduce overfitting given the modest sample size. Demographic predictors included country (Australia vs US), age (≥55 vs <55 years), and education (university vs non-university). Clinical predictors included country (Australia vs US), cancer stage (metastatic vs early-stage disease), treatment duration (≥12 vs <12 months), treatment interruption (yes vs no) and cardiometabolic comorbidity (yes vs no). Country was included in both models to account for differences between healthcare systems and survey administration. A complete case approach was used for regression analyses, with respondents with missing covariate data excluded. All analyses were considered exploratory.

## Results

### Participant demographic and clinical characteristics

A total of 75 individuals consented to participate. The total number of individuals approached or exposed to the survey was not systematically recorded due to the use of combined in-clinic and remote recruitment strategies; therefore, view and participant rates could not be calculated. Seventy-four respondents were included in the final analysis following exclusion of one Australian respondent who did not complete items beyond treatment interruption (completion rate, 98.7%). Item completion was high, with minimal missing data. All demographic and clinical characteristics were self-reported and were not independently verified by the research team. All respondents were female, including 24 from Australia and 50 from the US (**Table 1**). Most respondents (81.1%) were aged ≥45 years, with similar age distributions between Australia (83.3%) and US (80.0%; p = 0.748). Educational attainment was comparable across countries, with university-level education reported by 54.2% of Australian and 60.0% of US respondents (p = 0.802). No Australian respondents identified as Aboriginal and/or Torres Strait Islander. Among US respondents, 94.0% identified as non-Hispanic, 4.0% as Hispanic, and 2.0% preferred not to answer; most identified as White (78.0%), followed by Black or African American (18.0%).

**Table 1.** Characteristics of survey respondents by country (n = 74)

| **Characteristic** | **Australia (n = 24)**<br>**n (%)** | **US (n = 50)**<br>**n (%)** | **p-value** |
| --- | --- | --- | --- |
| **Age group** |  |  | 0.748 |
| 18-24 | 0 (0.0) | 0 (0.0) |  |
| 25-34 | 1 (4.2) | 2 (4.0) |  |
| 35-44 | 3 (12.5) | 8 (16.0) |  |
| 45-54 | 9 (37.5) | 11 (22.0) |  |
| 55-64 | 5 (20.8) | 13 (26.0) |  |
| 65+ | 6 (25.0) | 16 (32.0) |  |
| **Sex** |  |  | 1.000 |
| Female | 24 (100.0) | 50 (100.0) |  |
| Male | 0 (0.0) | 0 (0.0) |  |
| **Highest level of education** |  |  | 0.802 |
| Non-university | 11 (45.8) | 20 (40.0) |  |
| University | 13 (54.2) | 30 (60.0) |  |
| **Aboriginal and/or Torres Strait Islander status<sup>a</sup>** |  |  |  |
| Yes | 0 (0.0) | - |  |
| No | 24 (100.0) | - |  |
| Prefer not to answer | 0 (0.0) | - |  |
| **Hispanic ethnicity<sup>b</sup>** |  |  |  |
| Yes | - | 2 (4.0) |  |
| No | - | 47 (94.0) |  |
| Prefer not to answer | - | 1 (2.0) |  |
| **Race<sup>b</sup>** |  |  |  |
| White | - | 39 (78.0) |  |
| Black or African American | - | 9 (18.0) |  |
| Prefer not to answer | - | 2 (4.0) |  |
| **Cancer stage** |  |  | 0.004 |
| Early-stage | 16 (66.7) | 21 (42.0) |  |
| Metastatic | 6 (25.0) | 29 (58.0) |  |
| Unsure | 2 (8.3) | 0 (0.0) |  |
| **HER2-treatment duration** |  |  | 0.059 |
| About to start treatment | 2 (8.3) | 2 (4.0) |  |
| 0-3 months | 4 (16.7) | 4 (8.0) |  |
| 3-6 months | 8 (33.3) | 8 (16.0) |  |

| Characteristic | Australia (n = 24)<br>n (%) | US (n = 50)<br>n (%) | p-value |
| --- | --- | --- | --- |
| 6-9 months | 0 (0.0) | 2 (4.0) |  |
| 9-12 months | 5 (20.8) | 7 (14.0) |  |
| 12 months or more | 5 (20.8) | 27 (54.0) |  |
| **Treatment interruption** |  |  | 1.000 |
| Yes | 8 (33.3) | 15 (31.3) |  |
| No | 15 (62.5) | 32 (66.7) |  |
| Unsure | 1 (4.2) | 1 (2.1) |  |
| Missing | 0 | 2 |  |
| **Comorbidities<sup>c,d</sup>** |  |  |  |
| Diabetes | 0 (0.0) | 7 (14.0) | 0.088 |
| Chronic kidney disease | 0 (0.0) | 2 (4.0) | 1.000 |
| Hypertension | 5 (20.8) | 19 (38.0) | 0.188 |
| Hyperlipidaemia | 3 (12.5) | 12 (24.0) | 0.358 |
| Cardiovascular disease | 1 (4.2) | 5 (10.0) | 0.657 |
| Other comorbidities | 10 (41.7) | 18 (36.0) | 0.798 |
| No comorbidities | 12 (50.0) | 18 (36.0) | 0.314 |
| **Medications<sup>c,d</sup>** |  |  |  |
| ACE inhibitor | 2 (8.3) | 3 (6.0) | 0.657 |
| ARB | 2 (8.3) | 7 (14.0) | 0.709 |
| Beta-blocker | 1 (4.2) | 18 (36.0) | 0.004 |
| SGLT2 inhibitor | 0 (0.0) | 2 (4.0) | 1.000 |
| No cardiovascular medications | 17 (70.8) | 27 (54.0) | 0.210 |
| Unsure about cardiovascular medications | 2 (8.3) | 1 (2.0) | 0.244 |

Values are presented as n (%). P values were calculated using Fisher's exact test. Percentages may not sum to 100 due to rounding. All variables were self-reported.
Abbreviations: ACE = angiotensin-converting enzyme inhibitor; ARB = angiotensin II receptor blocker; SGLT2 = sodium-glucose cotransporter 2 inhibitor; US = United States.
<sup>a</sup> Aboriginal and/or Torres Strait Islander status was collected in the Australian cohort only.
<sup>b</sup> Hispanic ethnicity and race were collected in the United States cohort only.
<sup>c</sup> Multiple responses were allowed.
<sup>d</sup> Respondents reporting the presence of each condition or medication.

Clinical characteristics differed between countries (**Table 1**). Australian residents were more likely to report early-stage disease (Australia 66.7% vs US 42.0%), whereas metastatic disease was more common among US respondents (Australia 25.0% vs US 58.0%; p = 0.004). There was no statistically significant difference in duration of HER2-directed therapy between countries, although there was a non-significant trend toward longer treatment duration among US respondents (p = 0.059). Rates of treatment interruption were similar between countries (Australia 33.3% vs US 31.3%; p = 1.000). The prevalence of comorbidities was low overall, with no differences between countries. Use of cardiac medications was uncommon in both countries; however, beta-blocker use was more frequent among US respondents compared with Australian respondents (36.0% vs 4.2%; p = 0.004).

### Willingness to take cardioprotective medication against cardiotoxicity

Overall, 74.3% of participants reported being likely or very likely to take cardioprotective medication, with similar proportions in Australia (79.2%) and the US (72.0%) (p=0.831) (**Figure 1**, **Table 2**). Willingness did not differ by cancer stage, although a higher proportion of respondents with early-stage disease reported being likely or very likely to take cardioprotective medication compared with those with metastatic disease (81.1% vs 65.7%; p = 0.709) (**Supplementary Table 1).** A minority reported low willingness (10.8%), while 14.9% were neutral. Among participants reporting low willingness (n=8), concerns were heterogeneous and included dislike of additional medications and uncertainty regarding the necessity and benefit of cardioprotective therapy (**Supplementary Table 2**).

**Figure 1.**
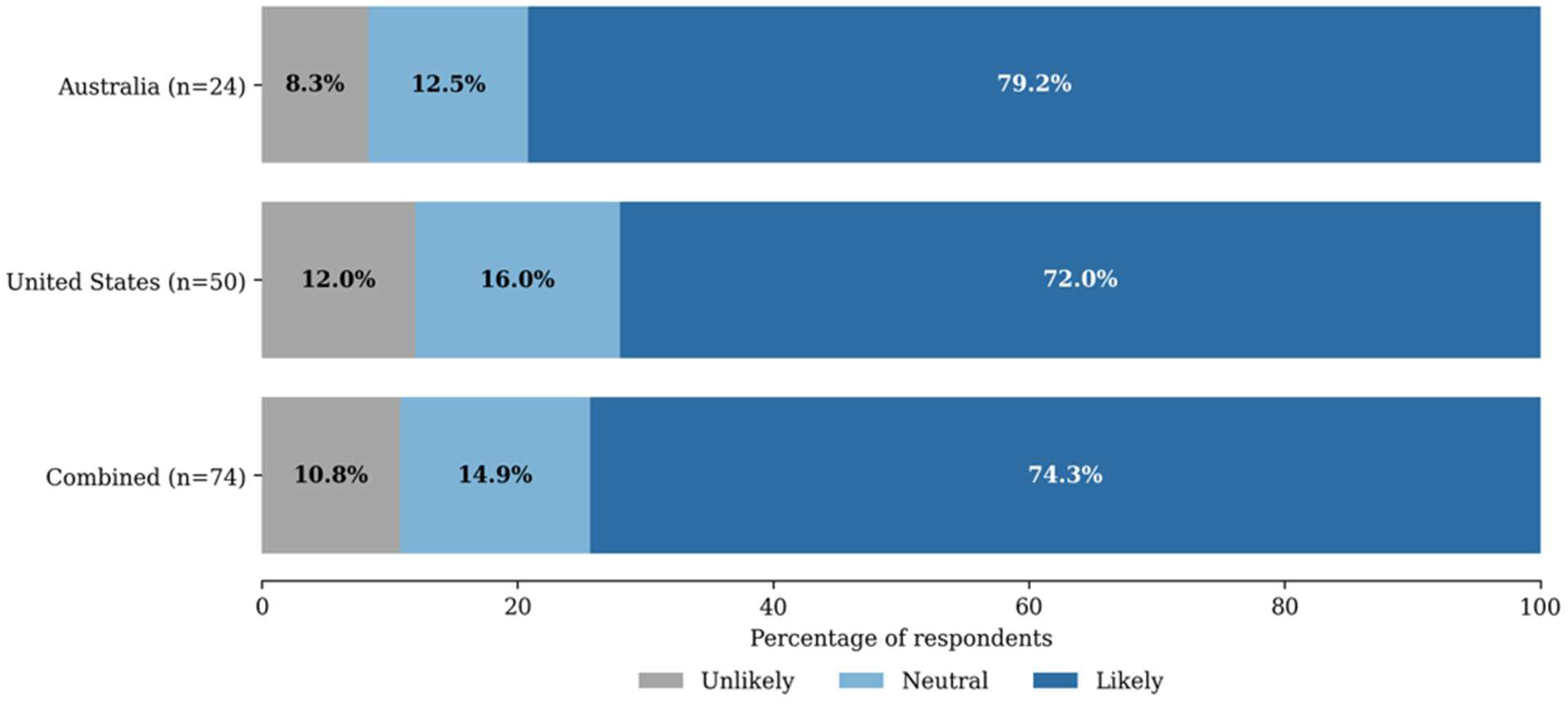
Willingness to take cardioprotective medication by country. Stacked bar chart showing willingness to take cardioprotective medication in Australia (n = 24), the United States (n = 50), and the combine sample (n = 74). Responses were collapsed into unlikely (very unlikely/unlikely), neutral, and likely (likely/very likely). Values represent percentages of respondents within each category.

**Table 2.** Willingness to take cardioprotective medication and treatment preferences (n = 74).

| **Characteristic** | **Australia (n = 24)**<br>**n (%)** | **US (n = 50)**<br>**n (%)** | **p-value** |
| --- | --- | --- | --- |
| **Willingness to take cardioprotective medication** |  |  | 0.831 |
| Very unlikely | 1 (4.2) | 4 (8.0) |  |
| Unlikely | 1 (4.2) | 2 (4.0) |  |
| Neutral | 3 (12.5) | 8 (16.0) |  |
| Likely | 9 (37.5) | 12 (24.0) |  |
| Very likely | 10 (41.7) | 24 (48.0) |  |
| **Acceptable number of tablets per day<sup>a</sup>** |  |  | 0.407 |
| 1 tablet | 4 (18.2) | 12 (27.3) |  |
| 2 or more tablets | 11 (50.0) | 14 (31.8) |  |
| Unsure | 7 (31.8) | 18 (40.9) |  |
| **Acceptable treatment duration<sup>b</sup>** |  |  | 0.317 |
| <6 months | 1 (4.5) | 1 (2.3) |  |
| 6-12 months | 0 (0.0) | 5 (11.6) |  |
| 1-2 years | 0 (0.0) | 1 (2.3) |  |
| >2 years but not indefinitely | 1 (4.5) | 0 (0.0) |  |
| As long as needed | 17 (77.3) | 29 (67.4) |  |
| Unsure | 3 (13.6) | 7 (16.3) |  |
| Missing | 0 | 1 |  |
| **Most important treatment outcome** |  |  | 0.003 |
| Cost<sup>c</sup> | 0 (0.0) | 2 (4.2) |  |
| Quality of life | 2 (8.3) | 15 (31.3) |  |
| Cardiac function | 0 (0.0) | 0 (0.0) |  |
| Minimizing side effects | 0 (0.0) | 0 (0.0) |  |
| Keeping cancer under control | 2 (8.3) | 12 (25.0) |  |
| Avoiding trastuzumab treatment interruption | 0 (0.0) | 0 (0.0) |  |
| Cancer free or preventing cancer recurrence | 18 (75.0) | 19 (39.6) |  |
| Other | 2 (8.3) | 0 (0.0) |  |
| Missing | 0 | 2 |  |
| **Interest in participating in a research study testing cardioprotective medication** |  |  | 0.376 |
| Yes | 11 (47.8) | 16 (34.0) |  |
| No | 5 (21.7) | 8 (17.0) |  |

| Characteristic | Australia (n =<br>24)<br>n (%) | US (n = 50)<br>n (%) | p-value |
| --- | --- | --- | --- |
| Unsure | 7 (30.4) | 23 (48.9) |  |
| Missing | 1 | 3 |  |

Values are presented as n (%). P values were calculated using Fisher's exact test. Percentages may not sum to 100 due to rounding.
Abbreviations: US = United States.
<sup>a</sup> Assessed only among participants indicating neutral, likely, or very likely willingness to take cardioprotective medication (Australia n = 22; US = 44).
<sup>b</sup> Assessed only among participants indicating neutral, likely, or very likely willingness to take cardioprotective medication, excluding missing response for this variable (Australia n = 22; US = 43).
<sup>c</sup> Cost was included as a response option in the US survey only.

### Beliefs regarding cardioprotective medication

Beliefs regarding cardioprotective medication reflected both strong facilitators and areas of uncertainty (**Figure 2; Supplementary Table 3**). Most participants agreed that physician (treating clinician) recommendation would influence their decision-making (79.1%) and that long-term cardiovascular benefit was important (72.9%). Perceptions of benefits and risks were more variable. Just over half of participants agreed that benefits would outweigh risks (54.2%), while 29.2% were neutral. Similarly, perceptions regarding side effects were mixed, with 43.7% agreeing that side effects may outweigh benefits, 31.2% neutral, and 25.0% disagreeing. Uncertainty was a consistent finding, with 60.4% of participants indicating they would require stronger evidence before initiating treatment and 39.6% expressing uncertainty regarding benefit. Concerns related to treatment burden were also evident, including perceptions that adding medication may be burdensome (20.8% agree) and concerns about polypharmacy (20.8% agree), although these were accompanied by substantial disagreement and neutral responses. These findings were supported by high levels of perceived acceptability of cardioprotective therapy during cancer treatment across domains including acceptability (76.0%), appropriateness (74.0%) and feasibility (72.0%) (**Supplementary Figure 1; Supplementary Table 4**).

**Figure 2.**
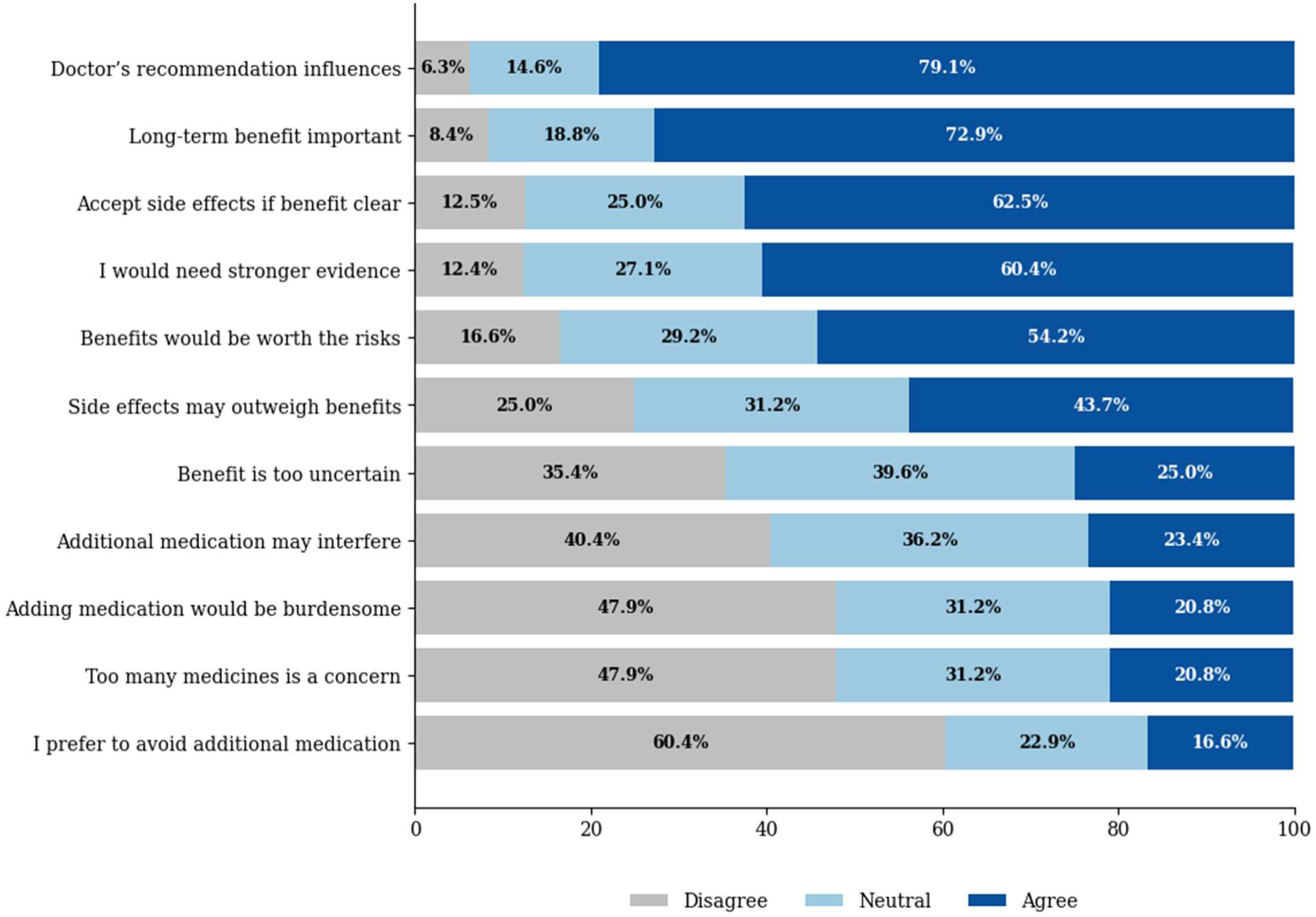
Beliefs regarding cardioprotective medication during cancer treatment (United State cohort; n = 50) Stacked bar chart showing responses to belief statements. Responses were collapsed into disagree (completely disagree/disagree), neutral, and agree (agree/completely agree). Values represent percentages of respondents within each category. Items are ordered by proportion agreeing.

### Preferences for cardioprotective medication

Respondents demonstrated flexibility regarding cardioprotective treatment preferences (**Table 2; Supplementary Table 1**). Approximately 38% of participants were willing to take two or more tablets per day, with no statistically significant difference between countries (Australia 50.0% vs US 31.8%; p = 0.407) or by cancer stage (early-stage 41.2% vs metastatic 30.0%; p = 0.357). The majority of respondents (70.8%) indicated willingness to continue treatment “as long as needed”, with similar proportions across countries (Australia 77.3% vs US 67.4%; p = 0.317) and no statistical difference by cancer stage (early-stage 81.8% vs metastatic 60.0%; p = 0.369). Preferences for treatment outcomes differed between countries (p = 0.003) (**Table 2)**. Cancer-related outcomes were most frequently prioritized, particularly prevention of cancer recurrence (Australia 75.0% vs US 39.6%). In contrast, US respondents more frequently prioritized quality of life (Australia 8.3% vs US 31.3%) and keeping cancer under control (Australia 8.3%, US 25.0%). Cardiovascular outcomes were not identified as the most important treatment outcome by participants in either country. In cancer stage-stratified analyses (**Supplementary Table 1**), preferences also differed by disease stage (p < 0.001). Prevention of cancer recurrence was the dominant priority among participants with early-stage disease (75.0% vs 26.5% in metastatic disease). Among respondents with metastatic disease, quality of life and keeping cancer under control were equally prioritized (both 35.3%).

Interest in participating in a research study testing cardioprotective medication was moderate and did not differ significantly between countries (Australia 47.8% vs US 34.0%; p = 0.376) or by cancer stage (early-stage 38.2% vs metastatic 35.3%; p = 0.903).

### Predictors of willingness to take additional cardioprotective medication

In multivariable analyses, no demographic or clinical variables were associated with willingness to take cardioprotective medication (**Figure 3**). A non-significant trend toward higher willingness was observed among respondents with cardiometabolic comorbidity (OR 3.20; 95% CI: 0.83–14.58). No associations were observed for age, education, cancer stage, treatment duration, or treatment interruption.

**Figure 3.**
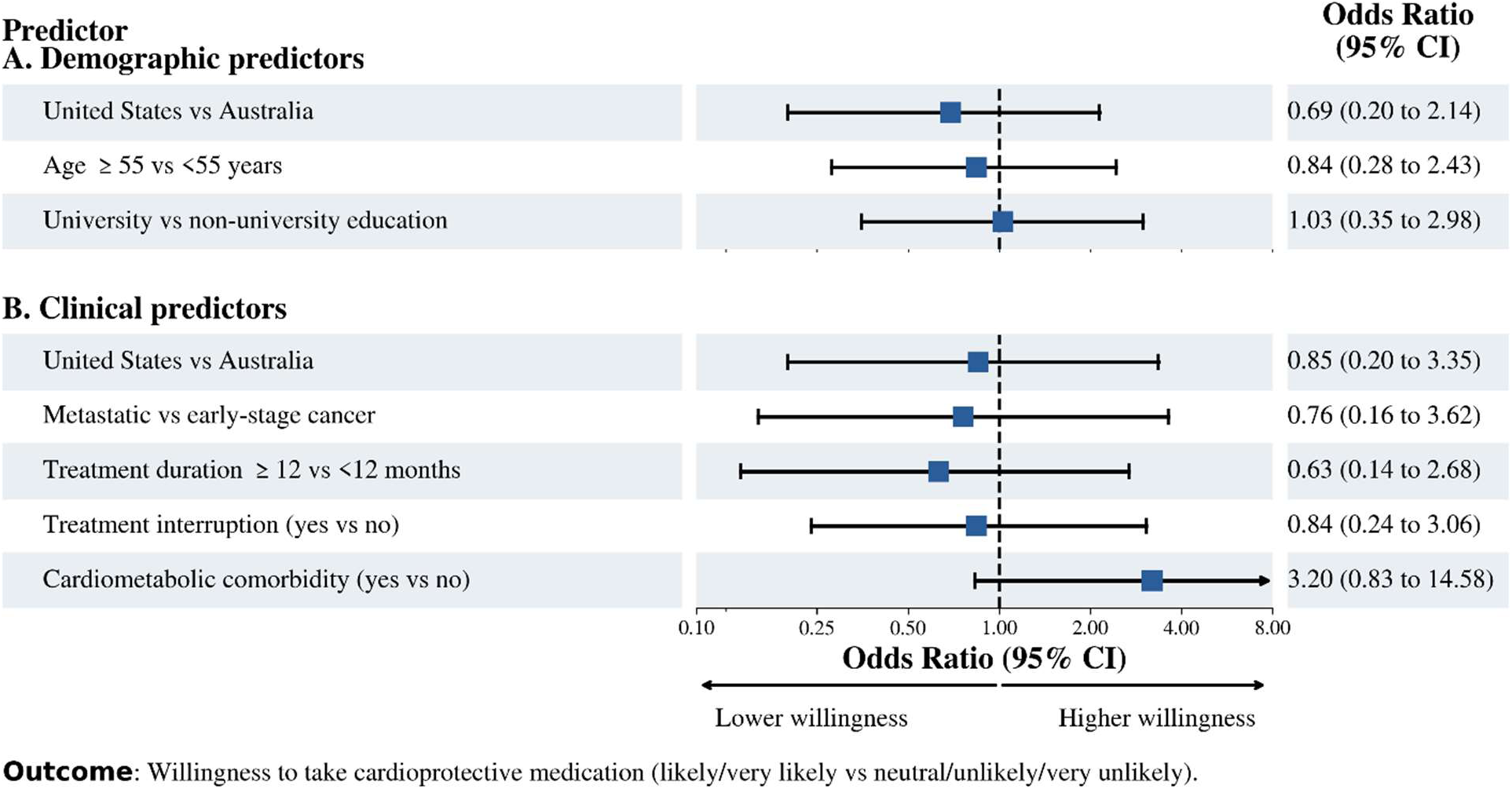
Predictors of willingness to take cardioprotective medications. Odds ratios (ORs) and 95% confidence intervals (CIs) for demographic (Model A; n = 74) and clinical (Model B; n = 69) predictors of willingness to take cardioprotective medication (likely/very likely vs neutral/unlikely/very unlikely). Squares represent point estimates and horizonal lines represent 95% CIs. The dashed vertical line indicates OR = 1. Values >1 indicate higher willingness. Arrows indicate truncated CIs extending beyond the axis range.

## Discussion

In this survey of individuals with HER2-positive breast cancer, approximately three-quarters of respondents reported willingness to take additional medication to prevent cardiotoxicity, and four-fifths indicated that a physician recommendation would be influential. While participants value long-term cardiovascular health, substantial uncertainty around benefit persists, with many indicating a need for stronger evidence before committing to an additional medication. Importantly, cardiovascular outcomes were not prioritized relative to cancer-related outcomes. Taken together, these findings suggest that willingness is not fixed but shaped by context, particularly how cardioprotective strategies are presented, including clinician endorsement and clear communication of evidence, and how these align with patients’ priorities during cancer treatment.

A central premise of cardiotoxicity prevention trials in patients receiving HER2 directed therapy is that any new intervention carries an implicit cost. Patients already managing a therapeutically demanding regimen are being asked to accept additional pharmacological burden when they have not yet developed cardiovascular side effects or disease. Unlike primary cardiovascular prevention in individuals without cancer, the context for cardio-oncology patients is active oncological treatment that may necessitate longitudinal and personalized cardiac surveillance.^16,17^ In this setting, patients’ willingness to participate in cardiotoxicity prevention trials in cardio-oncology is a prerequisite for the feasibility of cardiotoxicity prevention randomized trials, and our results are an encouraging signal for the field.

An important finding of this study concerns the motivation underlying patients’ willingness to accept additional medication for cardiotoxicity prevention. Cancer control remains the top priority. Patients frame their receptivity to cardioprotective medications through an oncologic lens which likely serves as an organizing principle of their clinical experience. And this willingness may evolve as treatment goals and perceptions of risk changed over the course of disease. This was reflected in stage-stratified findings, in which patients with early-stage disease predominantly prioritized prevention of cancer recurrence, whereas those with metastatic disease more frequently prioritized quality of life and maintaining cancer control, reflecting a shift toward disease control and symptom-focused outcomes in advanced disease. In addition, our findings suggest that physician recommendation may serve as an important facilitator of accepting such interventions, underscoring the importance of physicians’ familiarity with the evidence base and their ability to communicate that evidence in a way that aligns with patients’ priorities and motivations. It also reinforces the need for structured collaboration between oncology and cardiology to align cardiovascular risk and disease management decision-making with patients’ evolving cancer-related priorities.

The current landscape of cardiotoxicity prevention trials in patients receiving HER2-directed therapy is defined by heterogeneity in both design and findings. Many prior and ongoing trials have repurposed contemporary heart failure medications to prevent cardiotoxicity, yet signals remain mixed, due in part to small trial sizes and recruitment challenges.^12,18–24^ The proportion of screened patients ultimately randomized has ranged from 11 – 59% in key breast cancer cardiotoxicity prevention trials. The SCUSF 0806 trial only recruited 468 participants across 127 participating sites.^23^ Other non-breast cancer cardiotoxicity prevention trials face similar challenges. For example, the STOP-CA trial (Statins to Prevent the Cardiotoxicity of Anthracyclines) screened 1,600 patients over 4.5 years, of whom only 300 were randomized, with 46% of eligible patients declining participation.^25^ These recruitment difficulties reflect a broader reality. Patients with cancer frequently face competing priorities, balancing the demands of active cancer treatment with the added burden of research participation. Fundamental questions regarding the optimal dose and duration of cardioprotective strategies remain largely unexplored, and to date none of these trials have demonstrated a definitive effect on reducing cardiotoxicity.

In light of these challenges, our findings carry direct implications for future trial design, particularly strategies to optimize uptake, long-term adherence, and patient engagement. Although patients demonstrate high willingness (74%) to accept cardioprotective medications, this acceptance appears to be driven by cancer-related outcomes and influenced by physician recommendation and potential long-term benefits. Patients in our study also demonstrated flexibility regarding treatment burden, including willingness to take multiple medications and continue long-term, which has implications for intervention design. Importantly, willingness to accept medication may not translate into sustained adherence, even if proven efficacious. This is particularly relevant given that adherence to preventive cardiovascular medications is already suboptimal in the general population,^26,27^ and among patients with breast cancer who face challenges with adherence to oral endocrine therapies.^28^ Trials that fail to account for these dynamics risk poor adherence and limited real-world effectiveness, even if recruitment is successful.^29^ As the field continues to build the evidence-base, understanding patient perspectives must develop in parallel. Doing so will ensure that when effective therapies are identified, the groundwork for successful implementation is already in place. This type of work has received limited attention in cardio-oncology.

Our study has several limitations. First, the sample size is modest, limiting statistical power and precision of estimates. Second, the US and Australian surveys differed slightly in wording and scope, which may limit direct comparability between countries. Notably, the Australian survey framed cardioprotective medication as “proven to be safe and effective”, whereas the US survey used more uncertain language “may be proven”, which may have influenced reported willingness. Third, all data were self-reported and not independently verified, including information on medication use, comorbid conditions, disease stage, and may therefore be subject to recall and social desirability bias. As the survey was anonymous, it was not possible to verify or prevent multiple submissions; however, recruitment procedures were designed to minimize duplicate responses. All US participants were recruited in person, further reducing this risk. Additionally, recruitment across two countries (Australia and the US) somewhat strengthens the generalizability of findings. Fourth, a small number of US participants were recruited through cardio-oncology clinics, and these individuals may therefore be more attuned to cardiovascular health, although their inclusion does not appear to skew results toward greater acceptance of cardioprotective strategies. Fifth, the US survey was more expansive in scope, incorporating items designed to explore the motivations underlying patients’ willingness to accept additional medication to prevent cardiotoxicity. Survey length was balanced against participant burden, particularly among individuals undergoing active cancer treatment and the risks of participation fatigue and emotional harm. Finally, non-response bias cannot be excluded, as individuals who chose to participate may differ systematically from those who did not. Participants may not be fully representative of the broader population of patients with breast cancer, as respondents may be more health-literature or engaged with their care, and we had limited representation of ethnic diversity and did not capture socioeconomic or insurance-related factors that may influence access to and perceptions of preventative therapies.

## Conclusion

In conclusion, most patients with a history of HER2-positive breast cancer express willingness to accept cardioprotective medication, though this willingness is driven primarily by cancer-related outcomes. As the field works toward identifying proven cardioprotective strategies, these findings underscore the importance of patient-centered implementation research as parallel priorities.

## Data Availability

All data produced in the present study are available upon reasonable request to the authors.

## Abbreviations

HER2: human epidermal growth factor receptor
2 US: United States

## Competency in Medical Knowledge

Patients rely on physician guidance when considering cardioprotective medications that are highly acceptable, yet these recommendations must be framed in alignment with patients’ cancer related priorities.

## Translational Outlook

Future cardioprotective trials should be preceded by formative work characterizing patient perspectives to optimize trial design, recruitment strategies, and potential for real-world implementation.

## Central Illustration: Patient perspective on cardioprotective medication

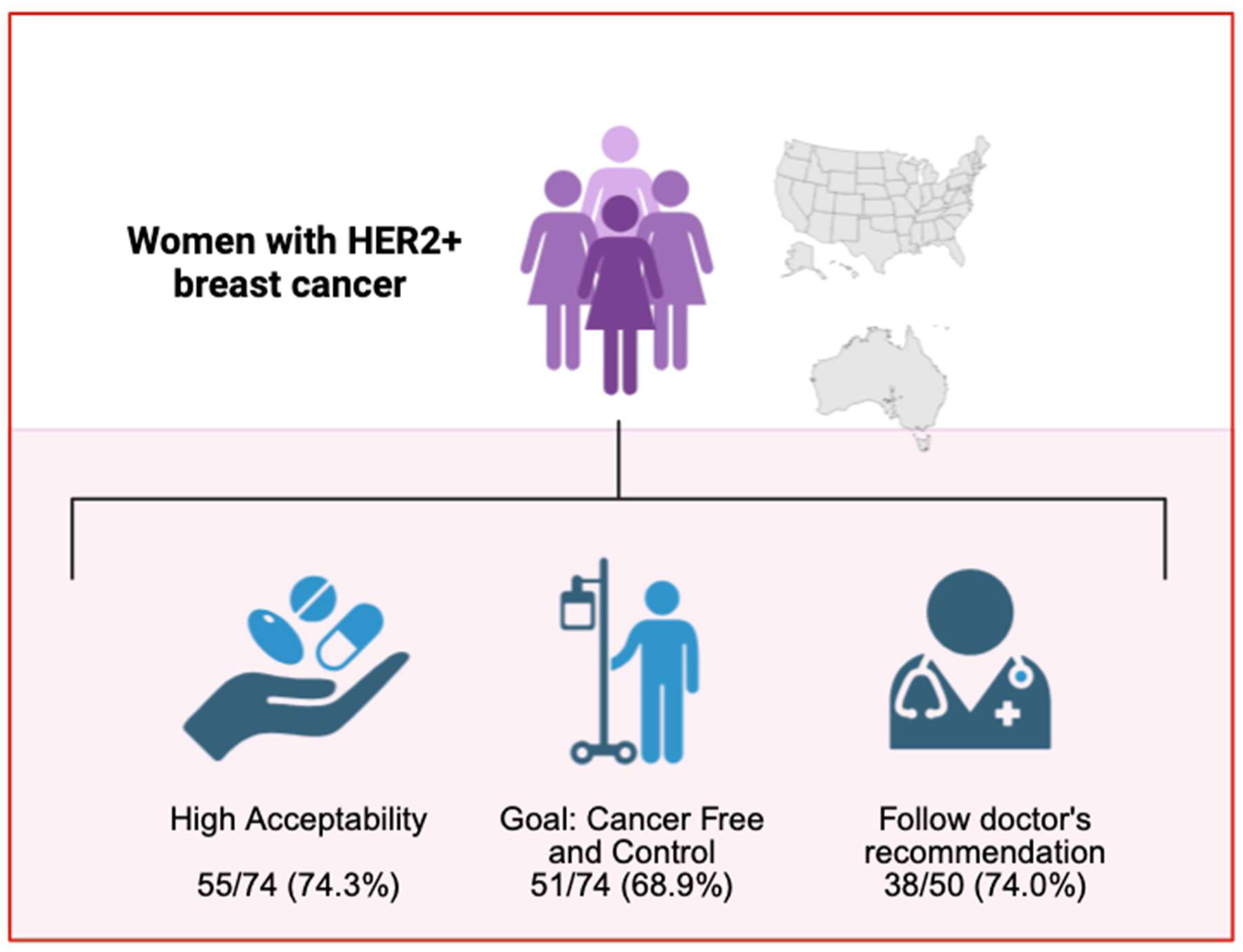

