## Supplementary Table 1 for "Patient perspectives on cardioprotective medication during breast cancer treatment"

**Supplementary Material**

**Supplementary Table 1. Willingness to take cardioprotective medication and treatment by cancer stage (n=72)[a]**

| Characteristic | Early-stage breast cancer (n = 37) n (%) | Metastatic breast cancer (n = 35) n (%) | p-value |
|---|---|---|---|
| **Willingness to take cardioprotective medication** | | | 0.709 |
| Very unlikely | 2 (5.4) | 3 (8.6) | |
| Unlikely | 1 (2.7) | 2 (5.7) | |
| Neutral | 4 (10.8) | 7 (20.0) | |
| Likely | 12 (32.4) | 9 (25.7) | |
| Very likely | 18 (48.6) | 14 (40.0) | |
| **Acceptable number of tablets per day[b]** | | | 0.357 |
| 1 tablet | 6 (17.6) | 10 (33.3) | |
| 2 or more tablets | 14 (41.2) | 9 (30.0) | |
| Unsure | 14 (41.2) | 11 (36.7) | |
| **Acceptable treatment duration[c]** | | | 0.369 |
| <6 months | 1 (3.0) | 1 (3.3) | |
| 6-12 months | 2 (6.1) | 3 (10.0) | |
| 1-2 years | 0 (0.0) | 1 (3.3) | |
| >2 years but not indefinitely | 0 (0.0) | 1 (3.3) | |
| As long as needed | 27 (81.8) | 18 (60.0) | |
| Unsure | 3 (9.1) | 6 (20.0) | |
| Missing | 1 | 0 | |
| **Most important treatment outcome** | | | <0.001 |
| Cost[d] | 1 (2.8) | 1 (2.9) | |
| Quality of life | 4 (11.1) | 12 (35.3) | |
| Cardiac function | 0 (0.0) | 0 (0.0) | |
| Minimizing side effects | 0 (0.0) | 0 (0.0) | |
| Keeping cancer under control | 2 (5.6) | 12 (35.3) | |
| Avoiding trastuzumab treatment interruption | 0 (0.0) | 0 (0.0) | |
| Cancer free or preventing cancer recurrence | 27 (75.0) | 9 (26.5) | |
| Other | 2 (5.6) | 0 (0.0) | |
| Missing | 1 | 1 | |

| Characteristic | Early-stage breast cancer (n = 37) n (%) | Metastatic breast cancer (n = 35) n (%) | p-value |
|---|---|---|---|
| **Interest in participating in a research study testing cardioprotective medication** | | | 0.903 |
| Yes | 13 (38.2) | 12 (35.3) | |
| No | 7 (20.6) | 6 (17.6) | |
| Unsure | 14 (41.2) | 16 (47.1) | |
| Missing | 3 | 1 | |

Values are presented as n (%). P values were calculated using Fisher's exact test. Percentages may not sum to 100 due to rounding.

[a] Excludes two respondents who reported 'Unsure' cancer stage.

[b]Assessed only among participants indicating neutral, likely, or very likely willingness to take cardioprotective medication (Early-stage n = 34; Metastatic = 30).

[c] Assessed only among participants indicating neutral, likely, or very likely willingness to take cardioprotective medication, excluding missing response for this variable (Early-stage n = 33; Metastatic = 30).

[d] Cost was included as a response option in the US survey only.
