## Supplementary Table 2 for "Patient perspectives on cardioprotective medication during breast cancer treatment"

**Supplementary Material**

**Supplementary Table 2. Concerns among respondents with low willingness to take cardioprotective medication (n=8)**

| Concerns | Australia (n = 2) n (%) | US (n = 6) n (%) |
|---|---|---|
| Distrust of medications | 0 (0.0) | 0 (0.0) |
| Dislike of taking additional medications | 1 (50.0) | 1 (16.7) |
| Cost | 0 (0.0) | 0 (0.0) |
| Side effects | 0 (0.0) | 0 (0.0) |
| Cardiotoxicity | 0 (0.0) | 1 (16.7) |
| Medication safety | 0 (0.0) | 0 (0.0) |
| Cardiotoxicity, but not sufficient to justify additional medication | 0 (0.0) | 1 (16.7) |
| Other | 1 (50) | 1 (16.7) |

Values are presented as n (%). Includes participants reporting unlikely or very unlikely willingness to take cardioprotective medication. Multiple responses were allowed.
Abbreviations: US = United States.
