## Supplementary Table 3 for "Patient perspectives on cardioprotective medication during breast cancer treatment"

**Supplementary Material**

**Supplementary Table 3. Beliefs about the benefits and risks of cardioprotective medication (United States cohort; n = 50)**

| Belief statement | Completely disagree n (%) | Disagree n (%) | Neither agree nor disagree n (%) | Agree n (%) | Completely agree n (%) | Missing n |
|---|---|---|---|---|---|---|
| Doctor's recommendation influences | 1 (2.1) | 2 (4.2) | 7 (14.6) | 22 (45.8) | 16 (33.3) | 2 |
| Long-term benefit important | 2 (4.2) | 2 (4.2) | 9 (18.8) | 19 (39.6) | 16 (33.3) | 2 |
| Accept side effects if benefit clear | 2 (4.2) | 4 (8.3) | 12 (25.0) | 22 (45.8) | 8 (16.7) | 2 |
| I would need stronger evidence before taking it | 3 (6.2) | 3 (6.2) | 13 (27.1) | 19 (39.6) | 10 (20.8) | 2 |
| Benefits would be worth the risks | 3 (6.2) | 5 (10.4) | 14 (29.2) | 20 (41.7) | 6 (12.5) | 2 |
| Side effects may outweigh benefits | 3 (6.2) | 9 (18.8) | 15 (31.2) | 17 (35.4) | 4 (8.3) | 2 |
| Benefit is too uncertain | 7 (14.6) | 10 (20.8) | 19 (39.6) | 8 (16.7) | 4 (8.3) | 2 |
| Additional medication | 8 (17.0) | 11 (23.4) | 17 (36.2) | 9 (19.1) | 2 (4.3) | 3 |

| **Belief statement** | **Completely disagree n (%)** | **Disagree n (%)** | **Neither agree nor disagree n (%)** | **Agree n (%)** | **Completely agree n (%)** | **Missing n** |
|---|---|---|---|---|---|---|
| may interfere with cancer treatment | | | | | | |
| Adding medication would be burdensome | 6 (12.5) | 17 (35.4) | 15 (31.2) | 7 (14.6) | 3 (6.2) | 2 |
| Too many medicines is a concern | 15 (31.2) | 8 (16.7) | 15 (31.2) | 5 (10.4) | 5 (10.4) | 2 |
| I prefer to avoid additional medication | 14 (29.2) | 15 (31.2) | 11 (22.9) | 5 (10.4) | 3 (6.2) | 2 |

Values are presented as n (%). Responses are showing across all Likert categories from completely disagree to completely agree. Percentages may not sum to 100 due to rounding. Items are ordered by proportion agreeing (agree or completely agree), with ties resolved using the proportion selecting “agree”. These data correspond to the collapsed categories in the Figure 2.
