## Supplementary Table 4 for "Patient perspectives on cardioprotective medication during breast cancer treatment"

**Supplementary Material**

**Supplementary Table 4. Acceptability of cardioprotective medication during cancer treatment (United States cohort; n = 50)**

| Items | Completely disagree n (%) | Disagree n (%) | Neither agree nor disagree n (%) | Agree n (%) | Completely agree n (%) | Missing n |
|---|---|---|---|---|---|---|
| Relevant | 3 (6.0) | 5 (10.0) | 4 (8.0) | 22 (44.0) | 16 (32.0) | 0 |
| Appropriate | 3 (6.0) | 5 (10.0) | 5 (10.0) | 20 (40.0) | 17 (34.0) | 0 |
| Manageable | 3 (6.0) | 5 (10.0) | 6 (12.0) | 18 (36.0) | 18 (36.0) | 0 |
| Acceptable | 4 (8.2) | 1 (2.0) | 10 (20.4) | 17 (34.7) | 17 (34.7) | 1 |
| Applicable | 3 (6.1) | 5 (10.2) | 7 (14.3) | 17 (34.7) | 17 (34.7) | 1 |
| Possible | 3 (6.0) | 5 (10.0) | 8 (16.0) | 19 (38.0) | 15 (30.0) | 0 |
| Welcome | 3 (6.0) | 4 (8.0) | 11 (22.0) | 15 (30.0) | 17 (34.0) | 0 |
| Easy | 3 (6.0) | 5 (10.0) | 11 (22.0) | 17 (34.0) | 14 (28.0) | 0 |
| Appealing | 3 (6.0) | 6 (12.0) | 10 (20.0) | 13 (26.0) | 18 (36.0) | 0 |

Values are presented as n (%). Responses are showing across all Likert categories from completely disagree to completely agree. Percentages may not sum to 100 due to rounding. Items are ordered by proportion agreeing (agree or completely agree), with ties resolved using the proportion selecting “agree”. These data correspond to the collapsed categories presented in Supplementary Figure 1.
