## Supplementary Figure 1 for "Patient perspectives on cardioprotective medication during breast cancer treatment"

**Supplementary Material**

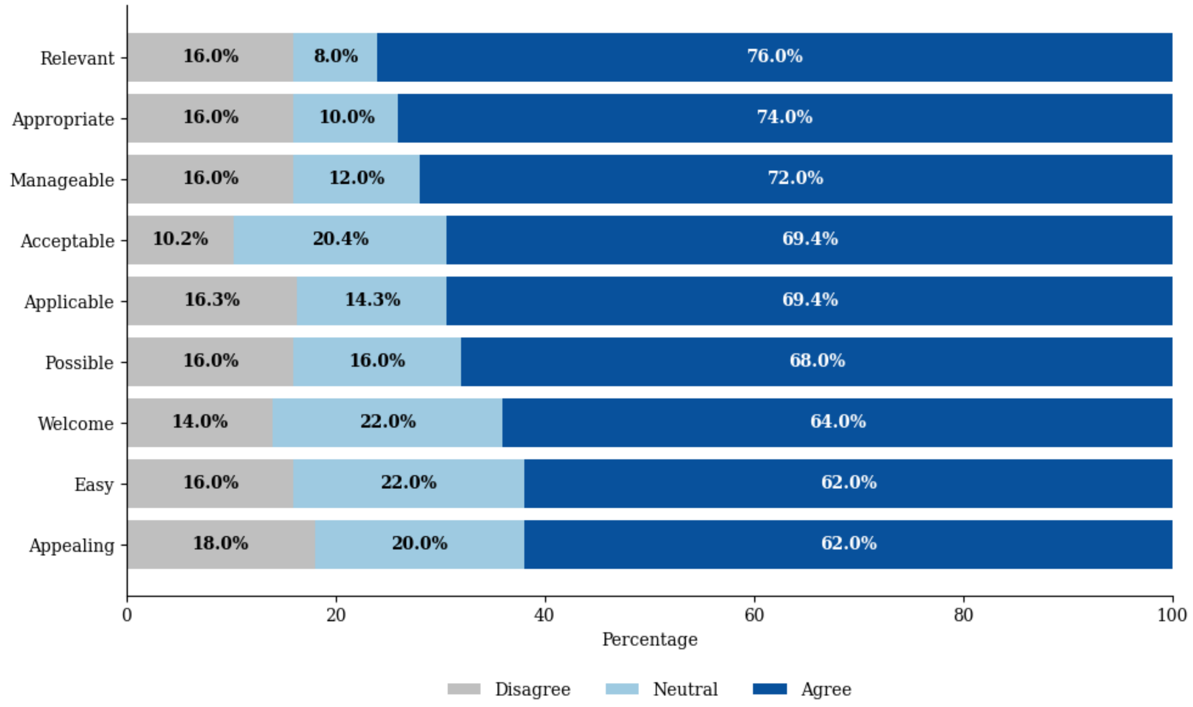


**Supplementary Figure 1. Acceptability of cardioprotective medication during cancer treatment (United States cohort; n = 50).**

Stacked bar chart showing participant responses to acceptability items. Responses were collapsed into disagree (completely disagree/disagree), neutral, and agree (agree/completely agree). Values represent percentages of respondents within each category. Items are ordered by proportion agreeing.
