## Supplementary Appendix 1 and 2 for "Patient perspectives on cardioprotective medication during breast cancer treatment"

### Supplementary Appendix 1 - Australian Survey Instrument

#### Participant demographics

**Please select the most appropriate response(s) from the range of options or type free text in the box provided for the following questions.**

What is your age? (Select one)

◯ 18 - 24 years old
◯ 25 - 34 years old
◯ 35 - 44 years old
◯ 45 -54 years old
◯ 55-64 years old
◯ 65 years or older

Sex recorded at birth (i.e. the sex on your original birth certificate) (Select one)

◯ Male
◯ Female
◯ Something else
◯ Prefer not to answer

Please specify:

__________________________________

What is your highest level of completed education? (Select one)

◯ Did not complete high school
◯ Completed high school
◯ Trade Certificate/Diploma (e.g. apprenticeship)
◯ Bachelor degree (including Honours)
◯ Masters degree
◯ Doctoral degree

Do you identify as being of Australian Aboriginal and/or Torres Strait Islander origin? (Select one)

◯ No
◯ Aboriginal
◯ Torres Strait Islander
◯ Both Aboriginal and Torres Strait Islander
◯ Prefer not to answer

#### Your clinical history

What is the stage of your breast cancer? (Select one)

Early-stage breast cancer has not spread beyond your breast or axillary lymph nodes. (This includes ductal carcinoma in situ and stage I, stage IIA and stage IIB).

Metastatic (or advanced) breast cancer has spread from the place of where it started to other places in your body. (This includes stage IIIA, stage IIIB, stage IIIC and stage IV).

◯ Early-stage
◯ Metastatic (or advanced) disease
◯ I don't know

How long did you receive trastuzumab / herceptin treatment for your breast cancer?

◯ I am about to start treatment
◯ 0 - 3 months
◯ 3 - 6 months
◯ 6 - 9 months
◯ 9 - 12 months
◯ 12 months or more

During your course of HER2+ breast cancer treatment, have you ever had to interrupt or stop your treatment prematurely?

◯ Yes
◯ No
◯ I don't know

If answered 'Yes', please explain why your treatment was interrupted or stopped.

In addition to your breast cancer diagnosis, are you currently diagnosed with any of the following health conditions? (Select all that apply)

☐ Diabetes
☐ Chronic kidney disease
☐ Hypertension (high blood pressure)
☐ Hyperlipidemia (high cholesterol)
☐ Cardiovascular disease (heart attack, angina, cardiac stent or bypass, stroke, peripheral vascular disease)
☐ Other health conditions
☐ No - I am only diagnosed with breast cancer

Please specify:

__________________________________

Do you take any of the following medications ? (Select all that apply)

☐ Angiotensin-converting enzyme (ACE) inhibitor (e.g. ramipril, enalapril, captopril, or perindopril)
☐ Angiotensin receptor blocker (ARBs) (e.g. telmisartan, olmesartan, candesartan, or losartan)
☐ Beta-blocker (e.g. bisoprolol, carvedilol or metoprolol)
☐ Sodium glucose cotransporter 2 (SGLT2) inhibitor? (e.g. empagliflozin or dapagliflozin)
☐ No
☐ I don't know

#### Willingness to take additional medication

For patients with HER2+ breast cancer, treatments that target the HER2 receptor have been shown to improve health and survival. But these medications also have potentially serious side effects in some patients.
The current management of HER2+ breast cancer includes a combination of standard chemotherapy with HER2-targeted antibody therapy (known as trastuzumab or herceptin). Adding trastuzumab to chemotherapy has led to a 37% improvement in overall survival.

Although trastuzumab is generally well tolerated, in 5 to 11 out of 100 (5% - 11%) of treated patients it can cause heart dysfunction - known as 'cardiotoxicity'. If cardiotoxicity occurs, you may need to interrupt or stop your trastuzumab to allow your heart to recover. Interrupting or stopping your trastuzumab may prevent you from achieving the full survival benefit offered by this treatment.

Taking additional heart medications may help to prevent this interruption. The side effects from these additional medications are minimal and may include the following: low blood pressure, dizziness, low blood sugar and minor infections.

---

How likely are you to take a medication (oral tablet or capsule) that has been proven to be safe and effective in minimizing your risk of cardiotoxicity during cancer treatment? (Select one)

◯ Very unlikely
◯ Unlikely
◯ Neutral
◯ Likely
◯ Very likely

---

How many oral tablets or capsules would you be willing to take daily? (Select one)

◯ 1
◯ 2
◯ I don't know

---

How long would you be prepared to take the medication(s) for? (Select one)

◯ Less than 6 months
◯ 6 months to 12 months (1 year)
◯ 1 to 2 years
◯ More than 2 years but not forever
◯ As long as needed to minimise my risk of developing heart disease
◯ I don't know

---

If you are concerned about taking medication(s) to minimise your risk of cardiotoxicity during cancer treatment, what are your concerns? (Select all that apply)

☐ I do not trust medications
☐ I do not like taking additional medications
☐ I am worried about the cost
☐ I am worried about the side effects
☐ I am worried about medication safety
☐ I am not concerned about cardiotoxicity
☐ I am concerned about cardiotoxicity but not enough to take additional medications
☐ Other

---

Please explain:

With respect to your breast cancer treatment, what is the most important outcome to you? (Select one)

◯ Quality of life
◯ Cardiac function
◯ Minimising side effects
◯ Keeping cancer under control
◯ Avoiding trastuzumab treatment interruption
◯ Cancer free or preventing cancer reoccurrence
◯ Other

Please explain

__________________________________

Would you be interested in participating in a research study testing medication(s) that may minimise your risk of cardiotoxicity during HER2+ breast cancer treatment?

◯ Yes
◯ No
◯ I don't know
