## Supplementary Appendix 1 and 2 for "Patient perspectives on cardioprotective medication during breast cancer treatment"

### Supplementary Appendix 2 – US Survey Instrument

#### Participant demographics

**Please select the most appropriate response(s) from the range of options or type free text in the box provided for the following questions.**

1) What is your age? (select one)

- ◯ 18 - 24 years old
- ◯ 25 - 34 years old
- ◯ 35 - 44 years old
- ◯ 45 - 54 years old
- ◯ 55 - 64 years old
- ◯ 65 years or older

2) Sex recorded at birth (i.e. the sex on your original birth certificate) (select one)

- ◯ Male
- ◯ Female
- ◯ Other - please specify:
- ◯ Prefer not to answer

3) What is your highest level of completed education? (select one)

- ◯ Did not complete high school
- ◯ Completed high school
- ◯ Bachelor degree
- ◯ Masters degree
- ◯ Doctoral degree

4) Are you of Hispanic, Latinx, or Spanish origin? (Select one)

- ◯ Yes
- ◯ No
- ◯ Prefer not to answer

5) How would you describe yourself? (Select one)

- ◯ American Indian or Alaska Native
- ◯ Asian
- ◯ Black or African American
- ◯ Native Hawaiian or Other Pacific Islander
- ◯ White
- ◯ Other
- ◯ Prefer not to answer

#### Your Medical History

6) What is/was the stage of your HER2+ breast cancer? (Select one)

Early-stage breast cancer has not spread beyond your breast or axillary lymph nodes. (This includes ductal carcinoma in situ and stage I, stage IIA and stage IIB).

Metastatic (or advanced) breast cancer has spread from the place of where it started to other places in your body. (This includes stage IIIA, stage IIIB, stage IIIC and stage IV).

- ◯ Early-stage
- ◯ Metastatic (or advanced) disease
- ◯ I don't know

7) How long did you receive trastuzumab/herceptin or cancer treatment directed at HER2 receptor for your breast cancer?

◯ I am about to start treatment
◯ 0 - 3 months
◯ >3 - 6 months
◯ >6 - 9 months
◯ >9 - 12 months
◯ >12 months or more

---

8) During your course of HER2+ breast cancer treatment, have you ever had to interrupt or stop your treatment prematurely?

◯ Yes
◯ No
◯ I don't know

---

If answered 'Yes' to question 8. Please explain why your treatment was interrupted or stopped.

__________________________________

---

9) In addition to your breast cancer diagnosis, are you currently diagnosed with any of the following health conditions? (Select all that apply)

☐ Diabetes
☐ Chronic kidney disease
☐ Hypertension (high blood pressure)
☐ Hyperlipidemia (high cholesterol)
☐ Cardiovascular disease (heart attack, angina, cardiac stent or bypass, stroke, peripheral vascular disease)
☐ Other health conditions - please specify:
☐ No - I am only diagnosed with breast cancer

---

10) Do you take any of the following medications? (Select all that apply)

☐ Angiotensin-converting enzyme (ACE) inhibitor (e.g. ramipril, enalapril, lisinopril, or perindopril)
☐ Angiotensin receptor blocker (ARBs) (e.g. telmisartan, olmesartan, candesartan, valsartan or losartan)
☐ Beta-blocker (e.g. bisoprolol, carvedilol, or metoprolol)
☐ Sodium glucose cotransporter 2 (SGLT2) inhibitor? (e.g. empagliflozin or dapagliflozin, or canagliflozin)
☐ No
☐ I don't know

---

Willingness to take additional medication

For patients with HER2+ breast cancer, treatments that target the HER2 receptor have been shown to improve health and survival. But these medications may also have potential side effects in some patients.
The current management of HER2+ breast cancer includes a combination of standard chemotherapy with HER2-targeted therapy (ie. trastuzumab).

Although trastuzumab or HER-2 targeted therapy is generally well tolerated, less than 5% (5 in 100) of treated patients it may cause heart dysfunction - known as 'cardiotoxicity'. If cardiotoxicity occurs, your doctor may need to interrupt or stop the trastuzumab to allow your heart to recover. Interrupting or stopping the trastuzumab may prevent you from achieving the full benefit offered by this treatment.

Currently, there are no treatments proven to prevent HER2 -related cardiotoxicity, although there is a growing interest in using other heart medications to reduce the risk of heart damage. These heart medications may help but we would like to explore your perspective on this approach. General side effects from these additional medications are minimal and may include the following: fatigue, low blood pressure, dizziness, low blood sugar and minor infections.

**11-13) The following questions will ask you about your willingness to take additional medication to lower your risk of cardiotoxicity during your breast cancer treatment.**

| | Completely disagree | Disagree | Neither agree nor disagree | Agree | Completely agree |
|---|---|---|---|---|---|
| Taking additional heart medication to reduce my risk of cardiotoxicity is acceptable to me. | ◯ | ◯ | ◯ | ◯ | ◯ |
| I find the idea of taking additional heart medication to reduce my risk of cardiotoxicity appealing | ◯ | ◯ | ◯ | ◯ | ◯ |
| I would welcome the additional heart medication to lower my risk of cardiotoxicity. | ◯ | ◯ | ◯ | ◯ | ◯ |
| Taking additional heart medication to lower my risk of cardiotoxicity seems relevant. | ◯ | ◯ | ◯ | ◯ | ◯ |
| Taking additional heart medication to lower my risk of cardiotoxicity seems like an appropriate option. | ◯ | ◯ | ◯ | ◯ | ◯ |
| Taking additional heart medication to lower my risk of cardiotoxicity seems applicable to me. | ◯ | ◯ | ◯ | ◯ | ◯ |
| Taking additional heart medication seems possible for me. | ◯ | ◯ | ◯ | ◯ | ◯ |
| Taking additional heart medications seems manageable. | ◯ | ◯ | ◯ | ◯ | ◯ |
| Taking additional heart medications seems easy. | ◯ | ◯ | ◯ | ◯ | ◯ |

14) How likely are you to take a medication (oral tablet or capsule) that may be proven to be safe and effective in minimizing your risk of cardiotoxicity during cancer treatment? (Select one)

◯ Very unlikely
◯ Unlikely
◯ Neutral
◯ Likely
◯ Very Likely

15) If you answered "Neutral," "Likely," or "Very likely" to question 14: How many additional tablets or capsules would you be willing to take daily? (Select one.)

◯ 1
◯ 2
◯ 3
◯ 4+
◯ I don't know

16) If you answered "Neutral," "Likely," or "Very likely" to question 14: How long would you be prepared to take the medication(s) for? (Select one)

- ◯ Less than 6 months
- ◯ 6 months to 12 months (1 year)
- ◯ 1 to 2 years
- ◯ More than 2 years but not forever
- ◯ As long as needed to minimize my risk of developing heart disease
- ◯ I don't know

17) If you answered "Very unlikely" or "Unlikely" to question 14: If you are concerned about taking medication(s) to minimize your risk of cardiotoxicity during cancer treatment, what are your concerns? (Select all that apply)

- ☐ I do not trust medications
- ☐ I do not like taking additional medications for prevention.
- ☐ I am worried about the cost of these additional medications.
- ☐ I am worried about the side effects of these medications.
- ☐ I am worried about medication safety
- ☐ I am not concerned about cardiotoxicity from cancer therapies.
- ☐ I am concerned about cardiotoxicity but not enough to take additional medications
- ☐ Other

If you answered "Other" to question 17, please explain

__________________________________

**18) Please indicate how much you agree or disagree with each of the following statements about the potential trade-offs involved in taking additional heart medication to reduce your risk of cardiotoxicity from your cancer therapy. Consider both the benefits and possible drawbacks when answering.**

| | Completely disagree | Disagree | Neither agree nor disagree | Agree | Completely agree |
|---|---|---|---|---|---|
| I am concerned that the side effects of additional heart medication may outweigh the benefits. | ◯ | ◯ | ◯ | ◯ | ◯ |
| I believe the benefit of reducing cardiotoxicity is worth the risk of possible side effects. | ◯ | ◯ | ◯ | ◯ | ◯ |
| I would accept some side effects if the medication significantly reduces my cardiotoxicity risk. | ◯ | ◯ | ◯ | ◯ | ◯ |
| I worry that taking additional heart medication would complicate my daily routine. | ◯ | ◯ | ◯ | ◯ | ◯ |
| I am willing to take more medications if it improves my long-term heart health. | ◯ | ◯ | ◯ | ◯ | ◯ |

| | | | | | |
|---|---|---|---|---|---|
| Managing multiple heart medications would be a burden for me, even if it reduced cardiotoxicity risk. | ◯ | ◯ | ◯ | ◯ | ◯ |
| I would prefer to avoid taking additional medication now, even if it increases my long-term risk. | ◯ | ◯ | ◯ | ◯ | ◯ |
| I am hesitant to take additional medication because the benefits are not guaranteed. | ◯ | ◯ | ◯ | ◯ | ◯ |
| I would be more likely to take additional medication if strongly recommended by my doctor. | ◯ | ◯ | ◯ | ◯ | ◯ |
| I would only consider additional heart medication if the benefits were clearly proven in people like me. | ◯ | ◯ | ◯ | ◯ | ◯ |
| I worry that additional heart medication may interfere with my current cancer treatment. | ◯ | ◯ | ◯ | ◯ | ◯ |

---

19) With respect to your breast cancer treatment, what is the most important outcome to you? (Select one)

◯ Quality of life
◯ Cardiac function
◯ Minimizing side effects
◯ Keeping cancer under control
◯ Avoiding trastuzumab treatment interruption
◯ Cancer free or preventing cancer reoccurrence
◯ Cost
◯ Other

---

If you answered "other" to question 19, please explain: ______________________________

---

20) If available, would you be interested in participating in a research study testing medication(s) that may minimize your risk of cardiotoxicity during HER2+ breast cancer treatment? ◯ Yes ◯ No ◯ I don't know
